# Nutritional awareness, eating behaviour, body mass index, and estimated resting metabolic rate in physical education students: A descriptive, analytical and correlational study

**DOI:** 10.64898/2026.10.06.26364876

**Authors:** Abdelnaser A. Qadumi, Rawn A. Qadumi, Ali A. Qadoume, Mahmoud Al-Atrash, Rawand K. Qutob, Sulaiman H. Amad, Mohammed A. Qadoumi, Qais M. Nairat, Abdelrahim K. Qaddoumi, Hala M. Jarrar

## Abstract

**Background:** University life can influence food choice even among physically active students. This study examined nutritional awareness, emotional, external, and restrained eating, body mass index (BMI), and estimated resting metabolic rate (RMR) in physical education students, and assessed differences by sex and academic level.

**Methods:** A descriptive, analytical and correlational design was used. The sample included 160 physical education students (95 women and 65 men; mean age 20.42 ± 1.53 years) from An-Najah National University. Nutritional awareness was assessed with a 15-item scale, and eating behaviour with the 33-item Dutch Eating Behavior Questionnaire. BMI was calculated from measured height and body mass, and RMR was estimated using an equation developed for athletes. Correlations, group comparisons, robust interaction models, and hierarchical regression were applied.

**Results:** Nutritional awareness was high (3.73 ± 0.81), whereas emotional (3.11 ± 1.18), external (3.20 ± 0.95), and restrained eating (3.25 ± 1.11) were moderate. Higher awareness was associated with less emotional (r = -0.617) and external eating (r = -0.652), but greater restrained eating (r = 0.644; all q < 0.001). These associations remained after adjustment. Men reported more external eating, while women reported more restrained eating. Eating patterns also varied by academic level. BMI and estimated RMR showed weaker and less consistent associations with eating behaviour.

**Conclusions:** Nutritional awareness was closely associated with reported eating behaviour, whereas BMI and estimated RMR were less informative. Sex and academic-level differences suggest that nutrition education for physical education students may be more useful when adapted to specific behavioural patterns. The findings represent associations and group differences and do not establish causal relationships.

## Introduction

Entering university gives students greater control over food choice, but those choices are made under practical constraints. Cost, time pressure, the campus food environment, academic demands, peer influence, and convenience all shape everyday eating [1,2]. Bayomy et al. studied students from health and non-health colleges at Northern Border University in Saudi Arabia and found unhealthy eating habits in more than 40% of each group; irregular meals, breakfast skipping, frequent fast-food intake, and low consumption of vegetables, fruits, and water were common [3]. Nutrition and lifestyle programmes can improve selected behaviours, although effects are not always sustained [4,5]. Moreover, knowledge alone rarely determines what students eat. In university samples, adjusted associations between nutrition literacy and dietary behaviour have generally been modest. Knowledge varies by sex and year of study, whereas dietary attitudes may show a closer relation to practice [6–8].

Physical education students warrant separate consideration because practical classes and sport occupy a substantial part of their routine. A high activity level, however, does not guarantee a nutritionally balanced diet. It is useful, therefore, to distinguish awareness of nutrition from reported eating patterns. Palestinian evidence links nutrition literacy with active information seeking and healthier food choices [9]. In male university rowers, a structured education programme also improved selected aspects of nutrition knowledge and dietary intake [10]. Together, these findings support nutrition education that is applied to the settings in which students study, train, and eat.

For this study, nutritional awareness referred to recognition of dietary practices relevant to health, training, and day-to-day food decisions. It was measured using the nutritional-awareness section of Qadumi’s Health Awareness Scale [11]. Because the items concern routine dietary practices, the resulting score was interpreted as practice-oriented awareness rather than a factual knowledge test. Eating behaviour was evaluated with the Dutch Eating Behavior Questionnaire (DEBQ), which assesses emotional, external, and restrained eating as distinct domains [12]. Studies in university samples support the instrument’s three-factor structure and its measurement invariance across sex [13,14]. Regional research also shows that diet, body weight, physical inactivity, sleep, and body-image concerns may cluster within the same individuals [15–18]. Accordingly, a high restrained-eating score was interpreted as greater cognitive restraint, not automatically as better eating.

BMI describes body mass relative to height. RMR is different: it estimates the energy needed to maintain basic physiological functions at rest. Body size and body composition are important determinants of RMR. If indirect calorimetry is unavailable, prediction equations are commonly used. Equations developed in athletic samples may improve estimation in active young adults, although error for an individual athlete can still be substantial [19,20]. Reviews of human energy expenditure also show that resting energy needs depend on several biological and body-composition factors [21]. For that reason, RMR should not be interpreted as a direct indicator of eating behaviour.

Findings on eating behaviour and body composition in university students have not been consistent [22]. Similar variation has been reported in regional adolescent studies, where dietary habits are linked with both physical activity and sedentary behaviour [23,24]. Stress is another factor, and emotional eating may rise during stressful periods [25]. Differences by sex are also reported, but their size and direction are not the same across studies [26]. Academic level may matter as well because students’ schedules, independence, and academic pressures change over time. Examining sex and academic level together may therefore reveal patterns that are missed in a simple overall comparison.

Adult studies provide additional context. Dietary preferences, meal habits, physical activity, and body composition often differ by sex [27–30]. Studies of adults with obesity also show that eating profiles can be related to body-composition patterns [31]. Comparative work across nutritional patterns reaches a similar conclusion [32]. These findings do not make BMI, RMR, or eating behaviour interchangeable. Rather, they show why these measures are better examined together than in isolation.

Most previous work has examined nutrition knowledge, eating behaviour, or anthropometric status separately. Much less is known about how nutritional awareness coexists with all three DEBQ domains when BMI and estimated RMR are considered in the same group of Palestinian physical education students. Studying these variables together may help separate behavioural correlates of awareness from anthropometric and metabolic correlates. It may also identify student subgroups that need different forms of nutrition support.

The study therefore had six objectives. We sought to: (1) describe nutritional awareness and emotional, external, and restrained eating; (2) describe BMI and estimated RMR; (3) examine the relationships between nutritional awareness and the three eating patterns; (4) examine how BMI and RMR relate to nutritional awareness and eating behaviour; (5) test differences by sex and academic level; and (6) examine sex-by-academic-level interactions. The descriptive, analytical, and correlational approach was selected to characterise these variables, examine their associations, and compare relevant student groups without making causal inferences.

## Materials and methods

### Study design and setting

The study population included 277 undergraduate physical education students enrolled during the second semester of 2025/2026. The final analytic sample contained 160 unique participants: 95 women (59.4%) and 65 men (40.6%). This represented 57.8% of the study population. No missing values were present in the variables used for the current analyses.

### Participants and power considerations

Eligible participants were undergraduate students enrolled in physical education. Participation was voluntary. The analysed sample was 18-25 years old and included students from all four academic levels. We did not have detailed information on non-participants. As a result, representativeness could not be judged beyond the overall participation fraction. This limitation is acknowledged rather than assumed away.

Sample size planning preceded the main analyses. For the primary correlation, G*Power 3.1 was specified for a two-sided alpha of 0.05, 80% power, and an anticipated correlation of r = 0.30. This yielded a minimum of 84 participants, well below the final sample of 160 [33]. We then examined the sensitivity of the 2 × 4 sex-by-academic-level interaction. With N = 160, approximately 80% power was available to detect an effect of f ≈ 0.264; smaller interactions may therefore have gone undetected.

### Anthropometric assessment

Anthropometric assessment was conducted in the Sports Sciences laboratory under a standardised morning protocol. Students attended between 08:00 and 10:00 before breakfast and were asked to avoid caloric intake for at least eight hours. Height and body mass were recorded with participants barefoot and wearing light clothing. Height was measured in centimetres and body mass in kilograms.

BMI was calculated as body mass in kilograms divided by height in meters squared (kg/m²). We classified values as underweight (<18.5 kg/m²), normal weight (18.5-24.9 kg/m²), overweight (25.0-29.9 kg/m²), or obesity (≥30.0 kg/m²).

### Estimated resting metabolic rate

RMR was not measured by indirect calorimetry. Instead, we estimated it with an equation developed for recreational athletes aged 18-35 years [19]: RMR (kcal/day) = 11.936 × body mass (kg) + 587.728 × height (m) - 8.129 × age (years) + 191.027 × sex + 29.279. Sex was coded 1 for men and 0 for women. In the original study, this equation performed better in active young adults than several equations designed for the general population. However, some error at the individual level is still expected [19,20].

### Nutritional awareness measure

We assessed nutritional awareness with the 15-item nutritional-awareness section of Qadumi’s Health Awareness Scale [11]. The items address common dietary practices, such as food variety, meal timing, food preparation, moderation of selected foods and drinks, and attention to nutrition information. Responses ranged from 1 (very low) to 5 (very high), and every item was scored in the same direction. The mean of the 15 responses was used as the participant’s score. Higher means indicated greater self-reported awareness. Because the questions are behaviour-oriented, we treated the score as awareness expressed through reported dietary practice rather than as factual nutrition knowledge. The original scale had strong internal consistency (Cronbach’s alpha = 0.91) [11]. Reliability in the present sample was also high (alpha = 0.979; McDonald’s omega = 0.979).

For descriptive purposes, the 1-5 scale was divided into five equal-width categories: very low (1.00-1.80), low (>1.80-2.60), moderate (>2.60-3.40), high (>3.40-4.20), and very high (>4.20-5.00). These bands describe score intensity only. They are not clinical cut-offs. Item-level findings are reported in the Results section.

### Dutch Eating Behavior Questionnaire

Eating behaviour was assessed with the 33-item DEBQ [12]. It includes 13 emotional-eating items, 10 external-eating items, and 10 restrained-eating items. Responses are given on a five-point scale. Each domain is scored separately because the three domains represent different constructs; a total DEBQ score is not recommended [12]. Higher domain scores indicate a stronger tendency toward that eating pattern.

The Arabic version used in the broader project was reviewed for language and content before the main study. Specialists in physical education evaluated the items, and pilot testing was completed. Earlier item-to-domain correlations in the project ranged from 0.44 to 0.85, indicating acceptable coherence within domains. Recent studies also support use of the DEBQ in university populations [13,14]. In the current sample, alpha/omega values were 0.965/0.966 for emotional eating, 0.897/0.898 for external eating, and 0.938/0.940 for restrained eating.

### Statistical analysis

The participant-level dataset was analysed in Python 3.13 using SciPy and statsmodels after variable definitions were checked against the source SPSS file. Continuous variables were summarized with means, standard deviations (SD), ranges, and 95% confidence intervals (CI). Categorical variables were reported as counts and percentages. Reliability was evaluated with Cronbach’s alpha and one-factor McDonald’s omega.

Pearson correlations were used for the prespecified association analyses because the scale distributions had acceptable skewness and kurtosis and the main interest was linear association. Fisher z 95% CIs were calculated. We controlled multiplicity across the 11 prespecified correlations with the Benjamini-Hochberg false discovery rate (FDR) procedure [34]. Spearman correlations were also examined as a sensitivity check. They did not materially change the three main relationships between nutritional awareness and eating behaviour.

Sex differences were examined with Welch’s independent-samples t test. This approach was used without assuming equal variances. We report mean differences, 95% CIs, and Hedges’ g. Academic-level differences were tested with Welch’s one-way ANOVA and summarized with the unequal-variance Cohen’s f. Significant omnibus tests were followed by Games-Howell comparisons. Sex-by-academic-level interactions were tested with ordinary least squares models using HC3 heteroskedasticity-consistent standard errors. Partial eta-squared from the corresponding factorial model is shown as a descriptive effect-size measure. Significant interactions were explored with Holm-adjusted simple sex contrasts within academic levels. All tests were two-sided with alpha = 0.05.

Estimated RMR is partly determined by body mass, height, age, and sex. We therefore added exploratory partial correlations adjusted for sex, age, and academic level to check whether the raw RMR-behaviour relationships were stable after basic demographic control. These analyses were treated as sensitivity checks, not replacements for the prespecified bivariate tests. Standardised anthropometric procedures and sensitivity analyses were used to reduce avoidable bias. However, the self-report scales remain vulnerable to social desirability and common-method bias. We also lacked detailed information on non-participants, so residual selection bias could not be quantified. Dietary intake misreporting procedures were not applicable because dietary intake itself was not measured.

We also fitted three hierarchical linear regression models as a sensitivity analysis. Emotional, external, and restrained eating were analysed as separate outcomes. Block 1 included sex, age, academic level, and BMI. Block 2 then added nutritional awareness. Academic level was entered as a categorical variable, with first year as the reference group. HC3 robust standard errors were used. We report standardised coefficients (β), robust 95% CIs, and the change in R² after nutritional awareness was added. FDR-adjusted q values were calculated across the three nutritional-awareness coefficients. The regressions were considered sensitivity analyses because the primary association objectives were defined as bivariate correlations.

### Use of generative artificial intelligence

Generative artificial intelligence was used only as an assistive tool for language editing and manuscript organization. The authors remained responsible for the study design, data, analysis, interpretation, and final wording.

### Ethics approval

The Institutional Review Board of An-Najah National University, Nablus, Palestine, approved the study on May 24, 2026 (Approval Reference No. Hum. ESci. May. 2026/33). All procedures followed the Declaration of Helsinki and relevant institutional requirements. Before participation, students received information about the study, confidentiality, voluntariness, and the right to withdraw. Written informed consent was obtained from every participant.

## Results

### Participant characteristics

As summarized in Table 1, the analytic sample included 160 students: 95 women (59.4%) and 65 men (40.6%). Mean age was 20.42 ± 1.53 years. Mean height was 168.41 ± 9.10 cm, and mean body mass was 65.79 ± 12.00 kg. BMI averaged 23.07 ± 2.93 kg/m², while estimated RMR averaged 1715.96 ± 255.22 kcal/day. Most students were in the normal BMI category (64.4%). A further 25.0% were overweight and 10.6% were underweight. No participant met the BMI threshold for obesity.

**Table 1.**
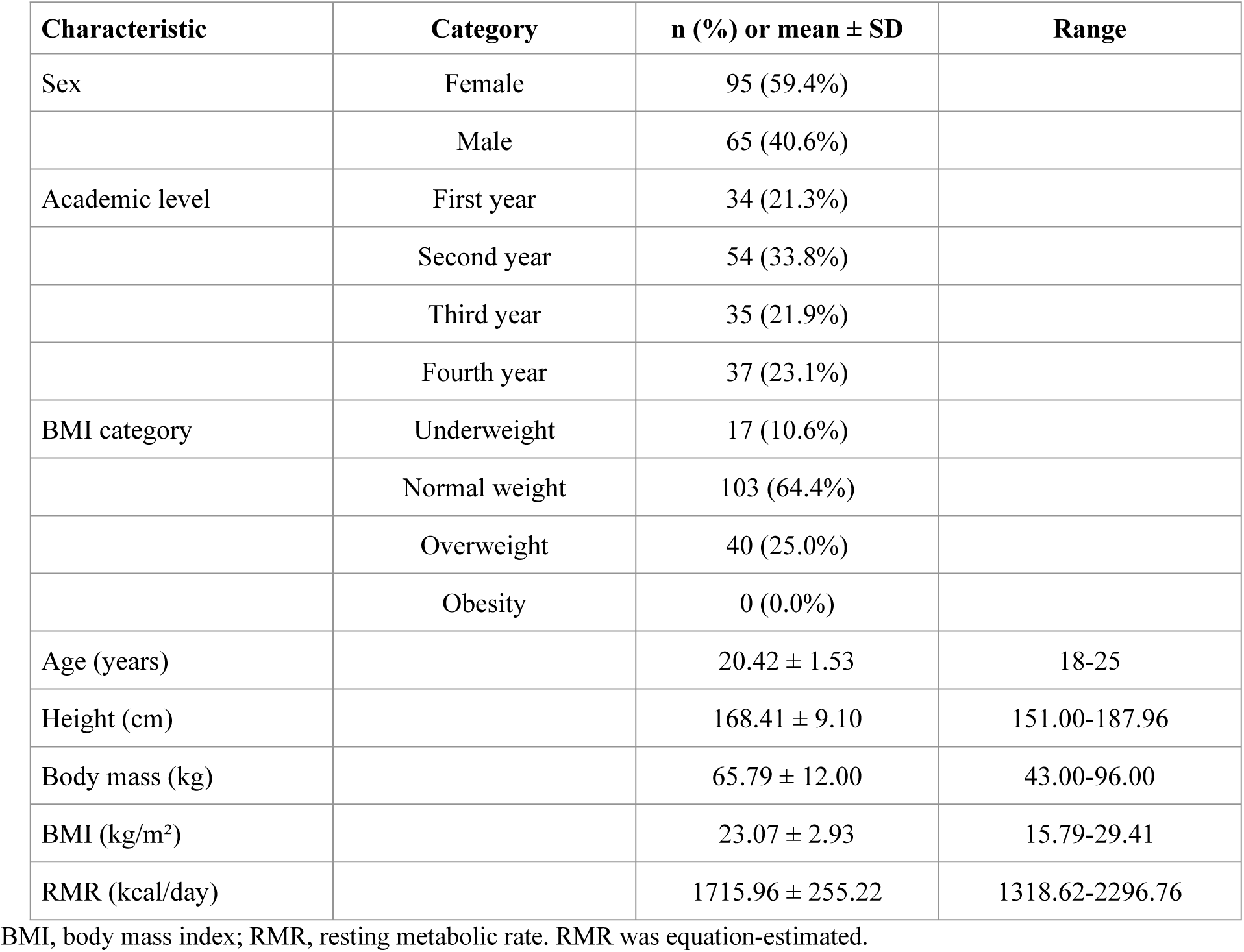
Participant characteristics (N = 160)

### Levels of nutritional awareness and eating behaviour

As detailed in Table 2, the highest nutritional-awareness mean was for maintaining an appropriate body weight (M = 4.30, SD = 1.04; 86.0%, very high). This was followed by eating at least three hours before training, competition, or practical classes (M = 4.24, SD = 1.09; 84.7%, very high) and varying the types of foods consumed (M = 4.21, SD = 1.08; 84.2%, very high). The lowest mean was for limiting pickles, spices, and pungent condiments (M = 2.67, SD = 1.49; 53.5%, moderate). Avoiding eating in the kitchen (M = 3.15, SD = 1.41; 63.0%, moderate) and reading calorie-content labels when buying food (M = 3.16, SD = 1.42; 63.1%, moderate) were also among the lowest-rated items. The high overall score therefore included clear variation across individual practices.

**Table 2.** Item-level descriptive statistics for nutritional awareness (N = 160)

| No. | Item | M | SD | Relative weight (%) | Level |
| --- | --- | --- | --- | --- | --- |
| 1 | I eat fresh fruits and vegetables more often than canned ones. | 4.07 | 1.19 | 81.4 | High |
| 2 | I maintain an appropriate body weight. | 4.30 | 1.04 | 86.0 | Very high |
| 3 | I vary the types of foods I consume. | 4.21 | 1.08 | 84.2 | Very high |
| 4 | I eat food at least three hours before training, competition, or practical classes. | 4.24 | 1.09 | 84.7 | Very high |
| 5 | I avoid eating in the kitchen. | 3.15 | 1.41 | 63.0 | Moderate |
| 6 | I drink limited amounts of cold water during training in hot weather. | 3.89 | 1.31 | 77.8 | High |
| 7 | I eat my meals at their scheduled times. | 3.84 | 1.31 | 76.9 | High |
| 8 | I prefer cooked food to fried food. | 4.12 | 1.24 | 82.4 | High |
| 9 | I eat balanced meals that include carbohydrates, fats, proteins, salts, minerals, vitamins, and water. | 4.03 | 1.27 | 80.5 | High |
| 10 | I consume pickles, spices, and pungent condiments in small amounts. | 2.67 | 1.49 | 53.5 | Moderate |
| 11 | I avoid eating meat before training, competition, or practical classes. | 3.81 | 1.28 | 76.2 | High |
| 12 | I avoid eating while listening to music or watching television. | 3.27 | 1.45 | 65.4 | Moderate |
| 13 | I largely avoid drinking tea and coffee. | 3.63 | 1.35 | 72.6 | High |
| 14 | I consume saturated (animal) fats in limited amounts. | 3.51 | 1.35 | 70.2 | High |
| 15 | I read the calorie-content label when purchasing food products. | 3.16 | 1.42 | 63.1 | Moderate |

Response range 1-5. Relative weight=(mean/5)×100. Descriptive categories: 1.00-1.80 very low; >1.80-2.60 low; >2.60-3.40
moderate; >3.40-4.20 high; >4.20-5.00 very high. Categories are descriptive, not clinical cut-offs.

Table 3 presents the item-level DEBQ results. Within emotional eating, boredom had the highest mean (M = 3.63, SD = 1.43; 72.5%, high), while fear had the lowest (M = 2.81, SD = 1.33; 56.1%, moderate). For external eating, the highest mean was shared by seeing delicious food and eating more when food tasted good (M = 3.65; 73.0%, high). The lowest mean was for difficulty resisting food simply because it was available (M = 2.84, SD = 1.32; 56.8%, moderate). In the restrained-eating domain, guilt after eating a large amount had the highest score (M = 3.41, SD = 1.42; 68.3%, high). Stopping before fullness had the lowest (M = 3.04, SD = 1.48; 60.9%, moderate). These item patterns identify the cues and weight-control thoughts that were most evident within each domain.

**Table 3.**
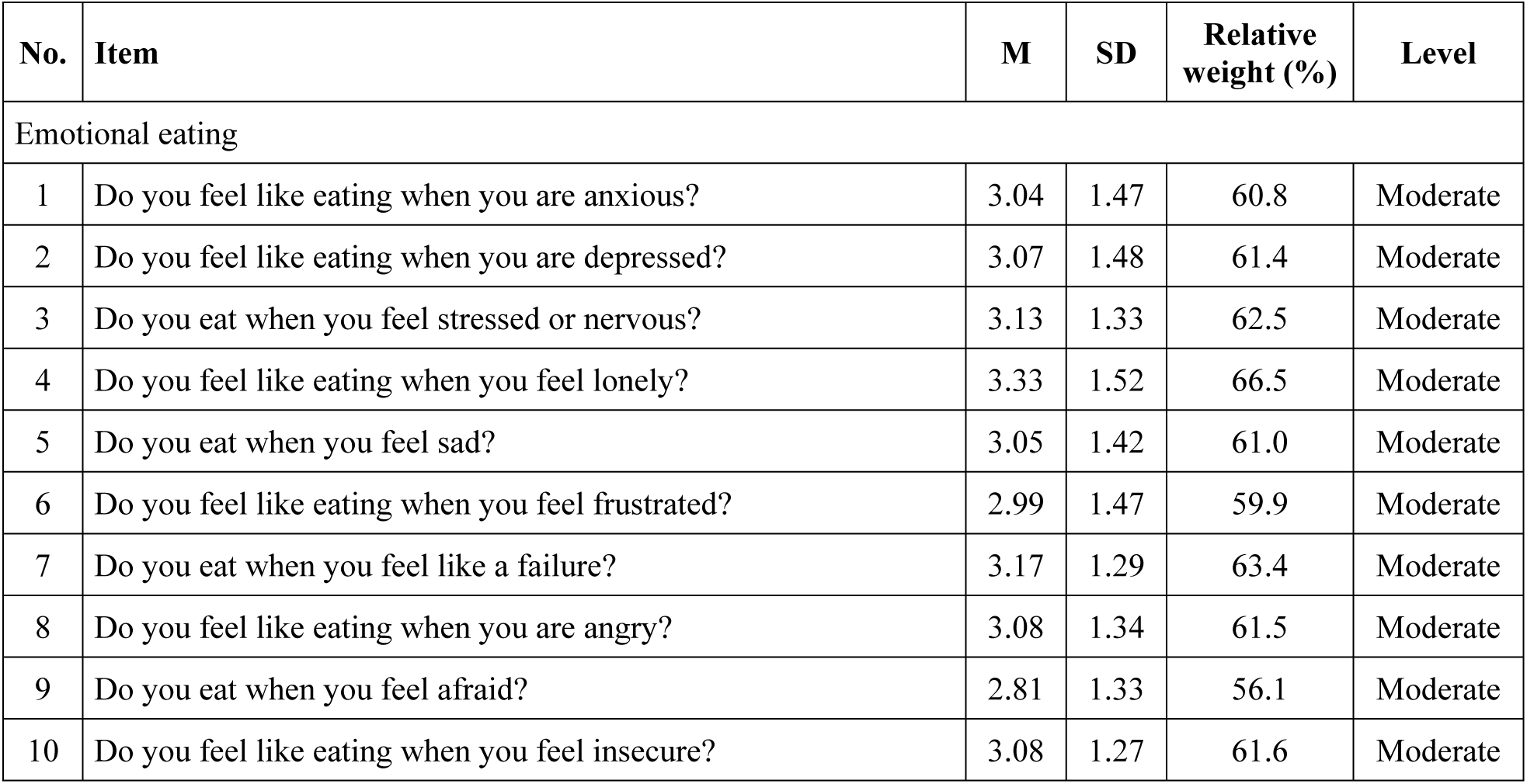
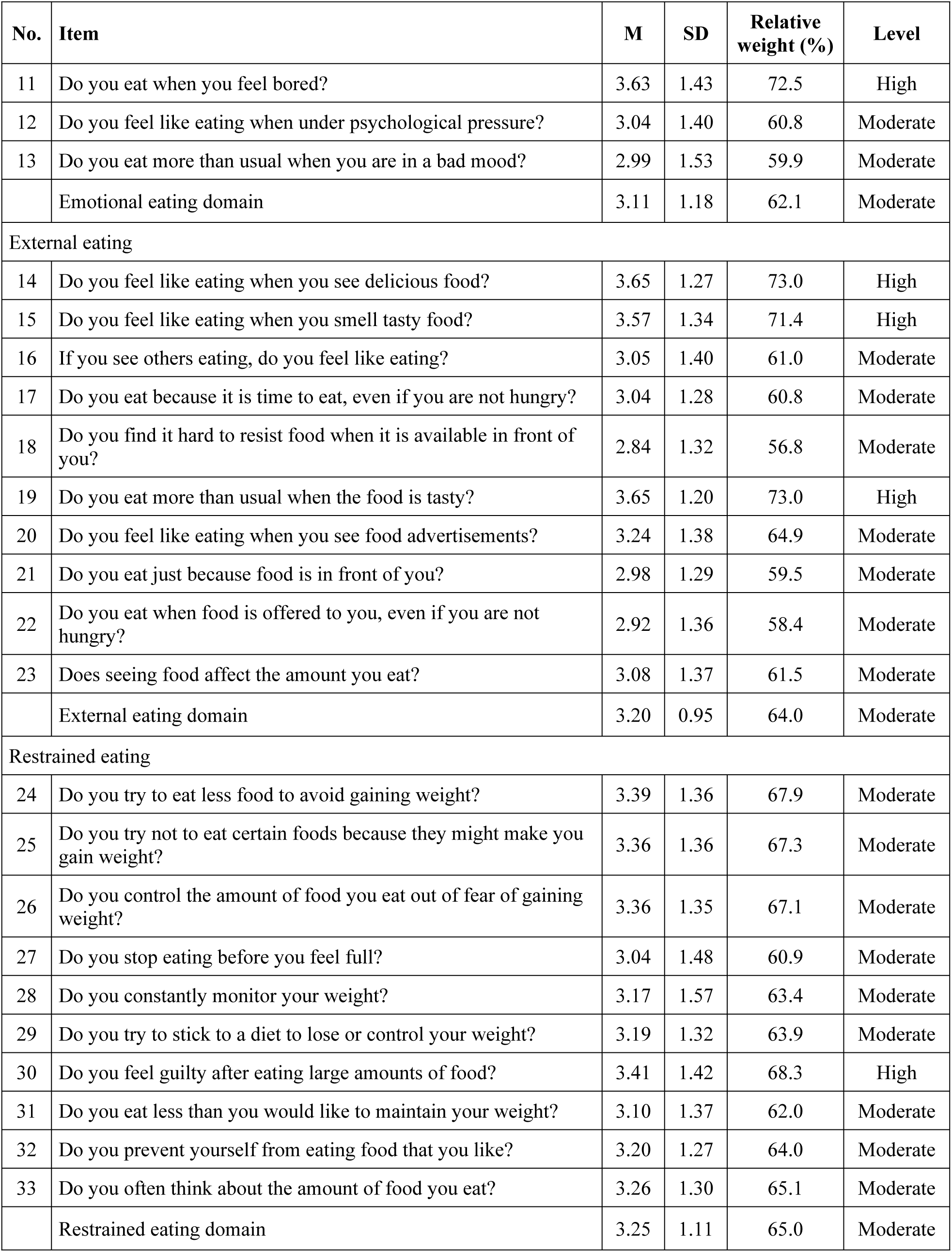
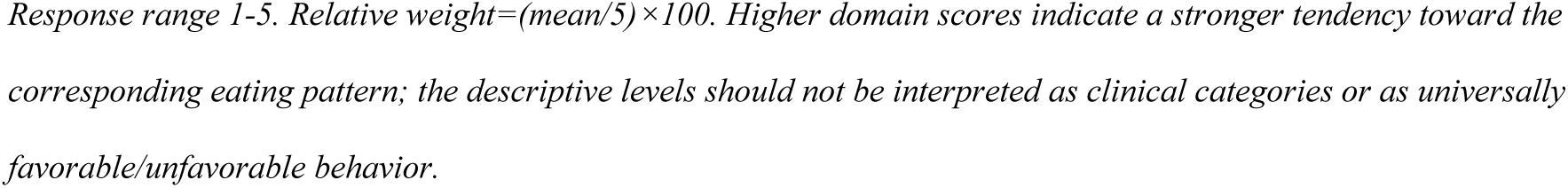
Item-level descriptive statistics for eating-behaviour patterns (N = 160)

Table 4 summarizes the scale-level descriptive statistics and reliability estimates. Mean nutritional awareness was 3.73 ± 0.81, corresponding to a relative weight of 74.5% and the high category. The three DEBQ domains were all in the moderate range: emotional eating, 3.11 ± 1.18; external eating, 3.20 ± 0.95; and restrained eating, 3.25 ± 1.11. Reliability was high or excellent for all four scores. As shown in Fig 1, nutritional awareness was the only mean that exceeded the threshold for the high category. The DEBQ values indicate the strength of each eating tendency. They should not be treated as simple ratings of healthy or unhealthy eating.

**Fig 1.**
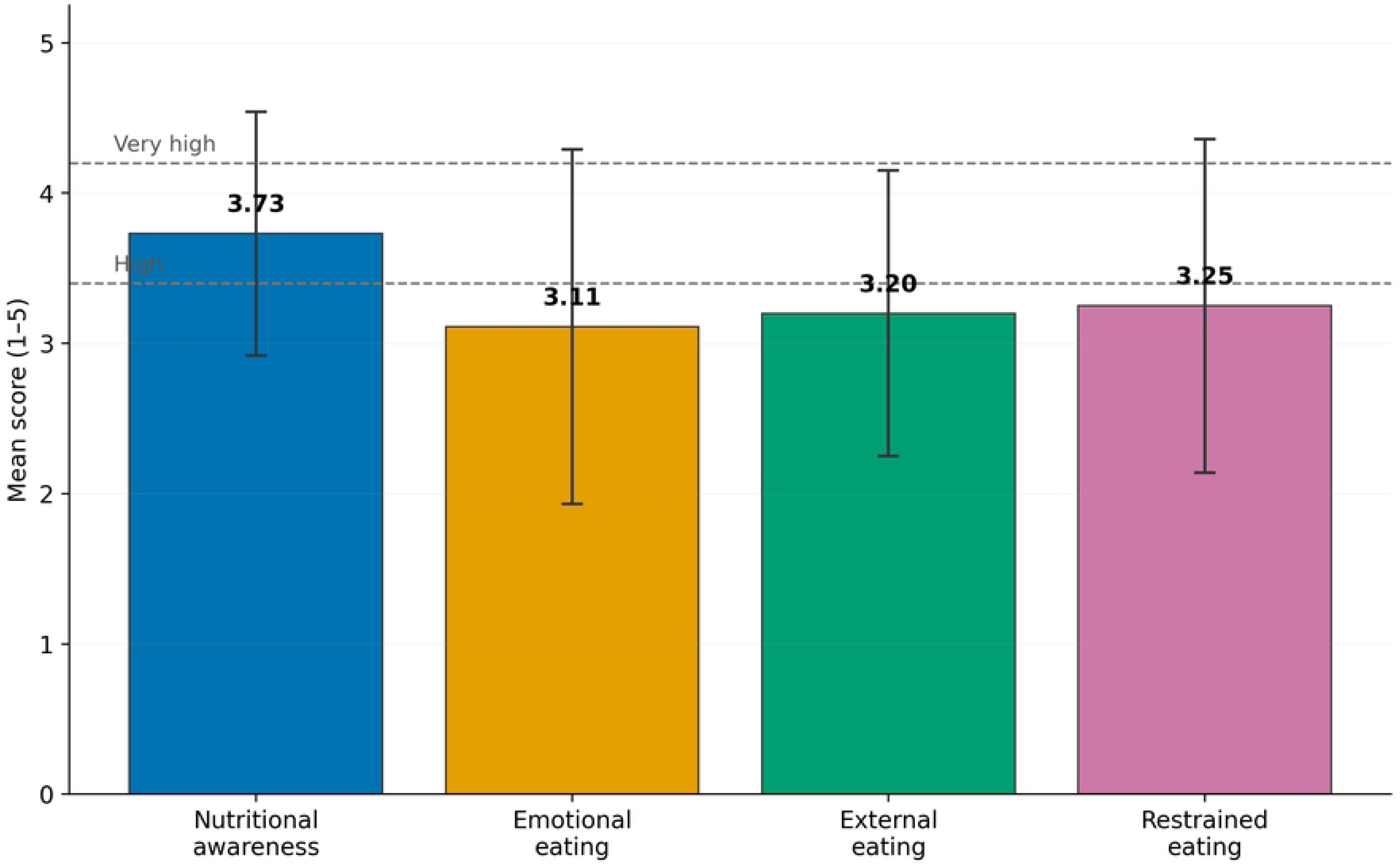
Mean levels of nutritional awareness and eating behaviour patterns. Error bars represent standard deviations. Dashed lines mark the high (>3.40) and very high (>4.20) descriptive thresholds.

**Table 4.** Descriptive statistics and reliability of study scales.

| Scale | M | SD | 95% CI | Relative weight (%) | Level | Alpha | Omega |
| --- | --- | --- | --- | --- | --- | --- | --- |
| Nutritional awareness | 3.73 | 0.81 | 3.60-3.85 | 74.5 | High | 0.979 | 0.979 |
| Emotional eating | 3.11 | 1.18 | 2.92-3.29 | 62.1 | Moderate | 0.965 | 0.966 |
| External eating | 3.20 | 0.95 | 3.05-3.35 | 64.0 | Moderate | 0.897 | 0.898 |
| Restrained eating | 3.25 | 1.11 | 3.08-3.42 | 65.0 | Moderate | 0.938 | 0.940 |

CI, confidence interval. Level categories are descriptive equal-width categories on the 1-5 response scale.

### Associations among nutritional awareness, BMI, RMR, and eating behaviour

As shown in Table 5, nutritional awareness was strongly related to all three eating patterns, but the directions differed. Higher awareness was associated with less emotional eating (r = -0.617, 95% CI -0.705 to -0.511, q < 0.001) and less external eating (r = -0.652, 95% CI -0.733 to -0.553, q < 0.001). In contrast, it was associated with greater restrained eating (r = 0.644, 95% CI 0.544 to 0.727, q < 0.001). The results therefore do not support treating eating behaviour as one simple ’healthier eating’ continuum.

**Table 5.** Prespecified bivariate associations.

| Variable 1 | Variable 2 | r | 95% CI | p | FDR q |
| --- | --- | --- | --- | --- | --- |
| Nutritional awareness | Emotional eating | -0.617 | -0.705 to -0.511 | <.001 | <.001 |
| Nutritional awareness | External eating | -0.652 | -0.733 to -0.553 | <.001 | <.001 |
| Nutritional awareness | Restrained eating | 0.644 | 0.544 to 0.727 | <.001 | <.001 |
| BMI | Nutritional awareness | -0.134 | -0.284 to 0.021 | 0.090 | 0.119 |
| BMI | Emotional eating | 0.169 | 0.014 to 0.316 | 0.032 | 0.051 |
| BMI | External eating | 0.195 | 0.041 to 0.339 | 0.014 | 0.025 |
| BMI | Restrained eating | 0.026 | -0.130 to 0.180 | 0.747 | 0.747 |
| RMR | Nutritional awareness | -0.131 | -0.281 to 0.024 | 0.098 | 0.119 |
| RMR | Emotional eating | 0.070 | -0.087 to 0.222 | 0.382 | 0.420 |
| RMR | External eating | 0.307 | 0.160 to 0.442 | <.001 | <.001 |
| RMR | Restrained eating | -0.200 | -0.345 to -0.046 | 0.011 | 0.025 |

Pearson correlations. FDR q values use the Benjamini-Hochberg procedure across the 11 prespecified correlations.

BMI was not significantly related to nutritional awareness or restrained eating. Its association with emotional eating was small and did not remain below the FDR threshold (r = 0.169, p = 0.032, q = 0.051). The BMI-external eating association was also small but retained FDR significance (r = 0.195, q = 0.025). In the unadjusted analyses, estimated RMR was positively related to external eating (r = 0.307, q < 0.001) and inversely related to restrained eating (r = -0.200, q = 0.025).

The hierarchical regression results are summarized in Table 6 and Fig 2. Covariates alone explained 12.7% of emotional eating, 19.5% of external eating, and 13.5% of restrained eating. After nutritional awareness was added, R² rose to 0.476, 0.558, and 0.511. The corresponding ΔR² values were 0.349, 0.363, and 0.376. Nutritional awareness remained strongly associated with emotional eating (adjusted β = -0.612, 95% CI -0.725 to -0.499, q < 0.001), external eating (β = -0.624, 95% CI -0.714 to -0.534, q < 0.001), and restrained eating (β = 0.636, 95% CI 0.529 to 0.742, q < 0.001). The main relationships were therefore not explained only by sex, age, academic level, or BMI.

**Fig 2.**
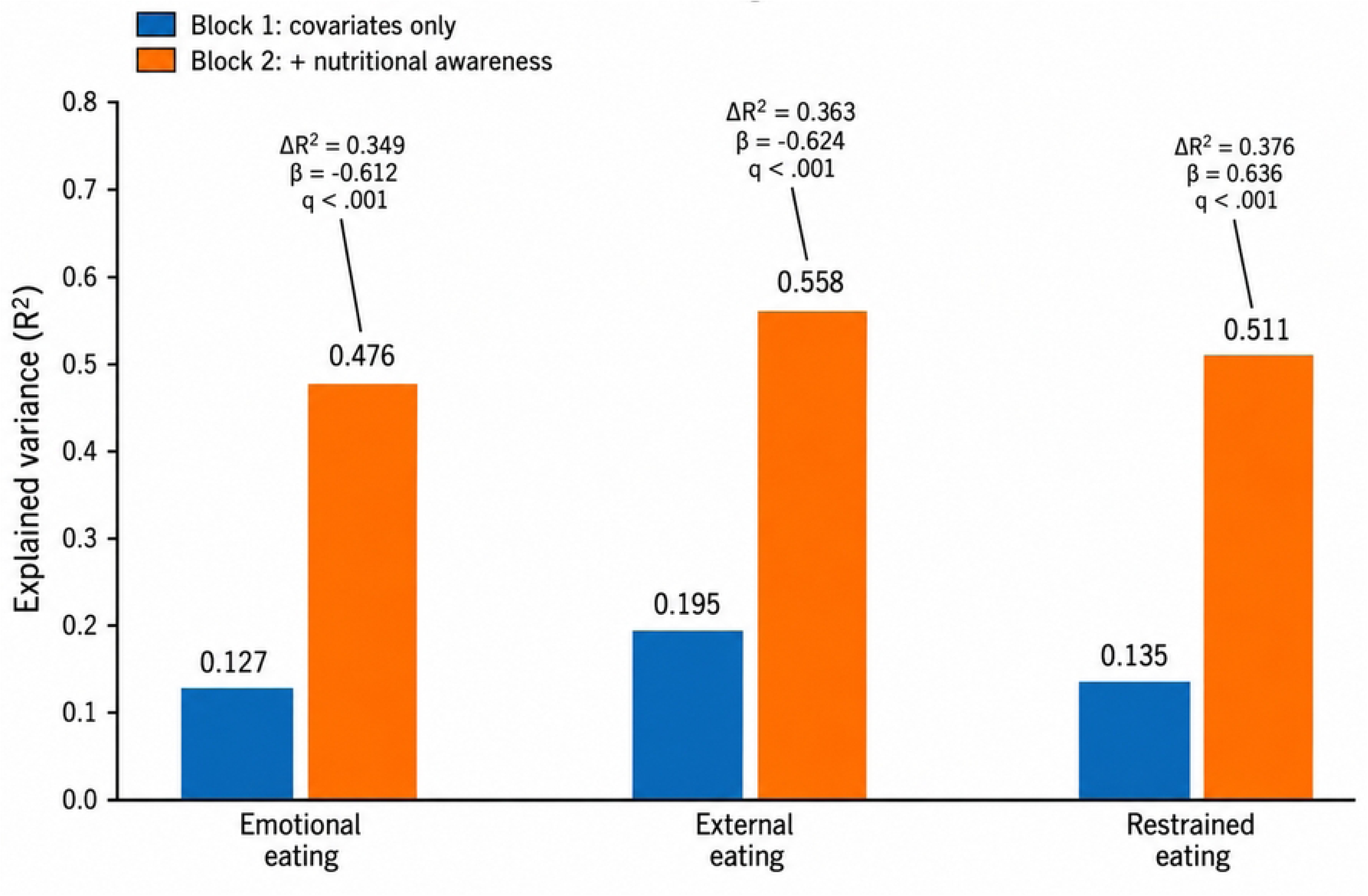
Hierarchical regression results for eating-behaviour outcomes. Bars show explained variance (R²) for Block 1 and Block 2. Block 1 included sex, age, academic level, and BMI; Block 2 added nutritional awareness. Labels above the Block 2 bars report ΔR², the standardised nutritional-awareness coefficient (β), and FDR q values.

**Table 6.** Hierarchical linear regression models for nutritional awareness and eating-behaviour outcomes.

| Outcome | Block 1 R<sup>2</sup> | Block 2 R<sup>2</sup> | $\Delta R^2$ | Adjusted $\beta$ | 95% CI | FDR q |
| --- | --- | --- | --- | --- | --- | --- |
| Emotional eating | 0.127 | 0.476 | 0.349 | -0.612 | -0.725 to -0.499 | <.001 |
| External eating | 0.195 | 0.558 | 0.363 | -0.624 | -0.714 to -0.534 | <.001 |
| Restrained eating | 0.135 | 0.511 | 0.376 | 0.636 | 0.529 to 0.742 | <.001 |

Block 1 included sex, age, academic level (categorical; first year as the reference category), and BMI. Block 2 added nutritional awareness. Adjusted $\beta$ is the standardised Block 2 coefficient with HC3 robust 95% CI; $\Delta R^2$ is the increase from Block 1 to Block 2. FDR q values use the Benjamini-Hochberg procedure across the three nutritional-awareness coefficients.

After adjustment for sex, age, and academic level, the raw RMR relationships with external and restrained eating were greatly reduced. The adjusted correlations were r = 0.015 (p = 0.848) and r = 0.082 (p = 0.300), respectively. This matters because the RMR equation itself includes sex and body-size variables. The full adjusted sensitivity results are presented in Table 7.

**Table 7.** Partial correlations adjusted for sex, age, and academic level.

| Exposure | Outcome | Partial r | p |
| --- | --- | --- | --- |
| BMI | Nutritional awareness | -0.077 | 0.335 |
| BMI | Emotional eating | 0.160 | 0.044 |
| BMI | External eating | 0.129 | 0.105 |
| BMI | Restrained eating | 0.106 | 0.183 |
| Estimated RMR | Nutritional awareness | 0.019 | 0.816 |
| Estimated RMR | Emotional eating | 0.165 | 0.037 |
| Estimated RMR | External eating | 0.015 | 0.848 |
| Estimated RMR | Restrained eating | 0.082 | 0.300 |

*Estimated RMR is mathematically derived from body mass, height, age, and sex; therefore, adjusted RMR analyses are explicitly exploratory.*

### Differences according to sex

As shown in Table 8, Welch’s tests found no significant sex difference in nutritional awareness or emotional eating. Men had higher external-eating scores than women (3.60 ± 1.03 vs 2.93 ± 0.80). The mean difference was 0.67 (95% CI 0.37 to 0.97; p < 0.001; g = 0.74). Women, however, had higher restrained-eating scores (3.50 ± 1.09 vs 2.89 ± 1.05). The male-minus-female difference was -0.61 (95% CI -0.95 to -0.27; p < 0.001; g = -0.57). Fig 3 displays these differences with SD error bars.

**Fig 3.**
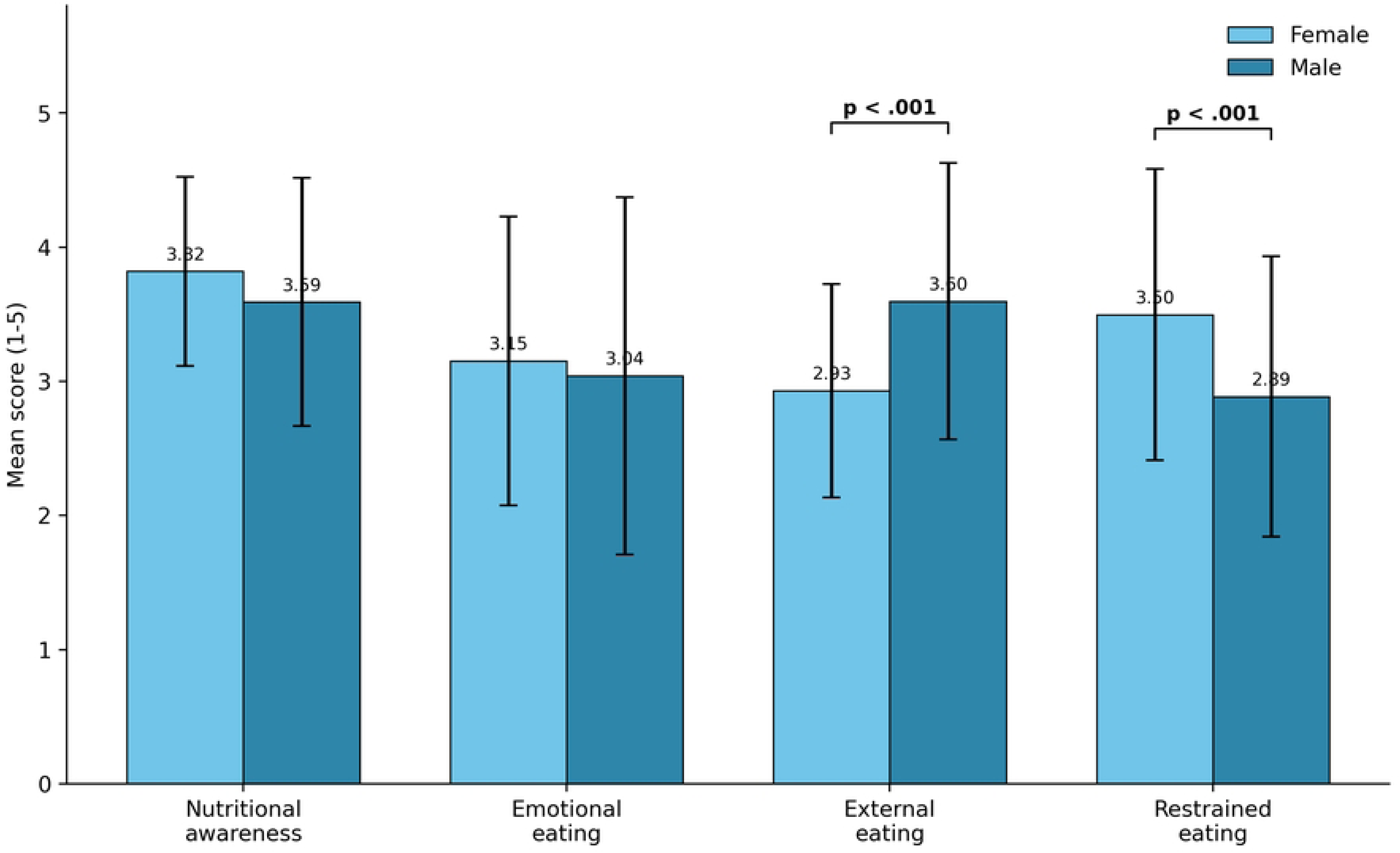
Sex differences in nutritional awareness and eating behaviour. Bars show means and SD error bars. Significance brackets identify Welch-test differences for external and restrained eating.

**Table 8.**
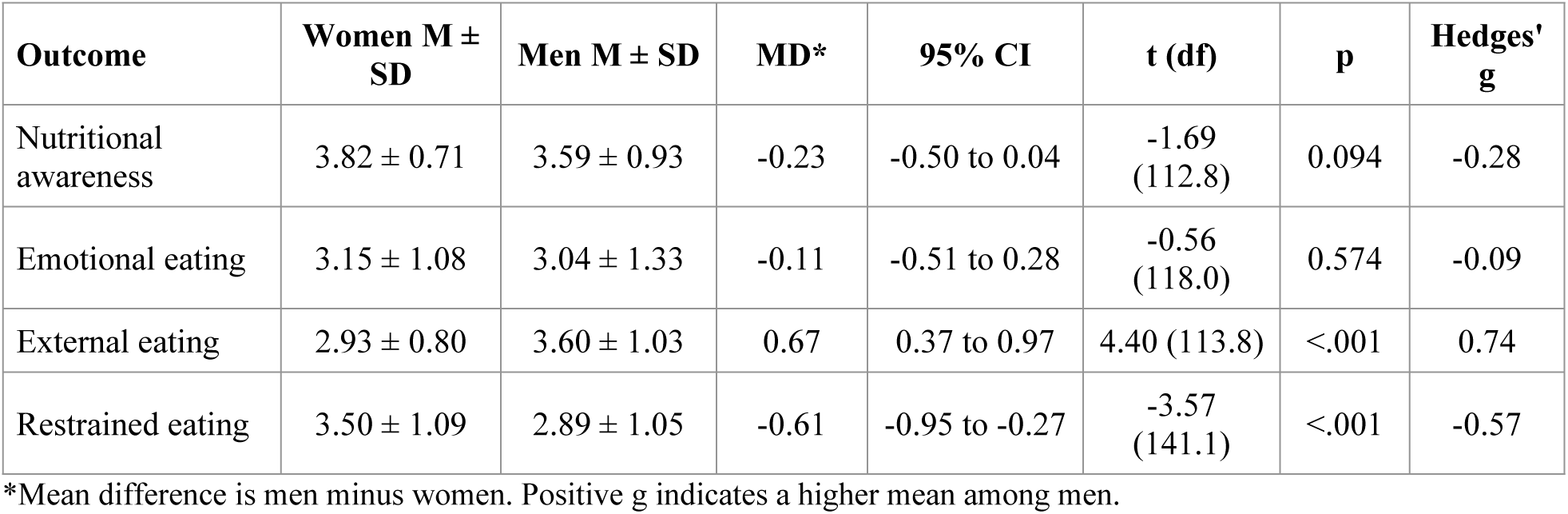
Welch sex comparisons for nutritional awareness and eating behaviour.

### Academic-level differences and sex-by-academic-level interactions

As summarized in Table 9, academic level was related to all three eating-behaviour domains, but not to nutritional awareness in the Welch omnibus analysis. Emotional eating differed by level (p = 0.002). First-year students scored higher than third-year students (3.48 vs 2.55; Games-Howell p = 0.007), and fourth-year students also scored higher than third-year students (3.43 vs 2.55; p = 0.006). External eating also differed (p = 0.011). First-year students scored higher than second-year students (3.63 vs 3.00; p = 0.016) and third-year students (3.63 vs 2.98; p = 0.019). Restrained eating differed as well (p = 0.018), with second-year students scoring higher than fourth-year students (3.57 vs 2.90; p = 0.024). The significant Games-Howell pairwise comparisons are listed in Table 10.

**Table 9.**
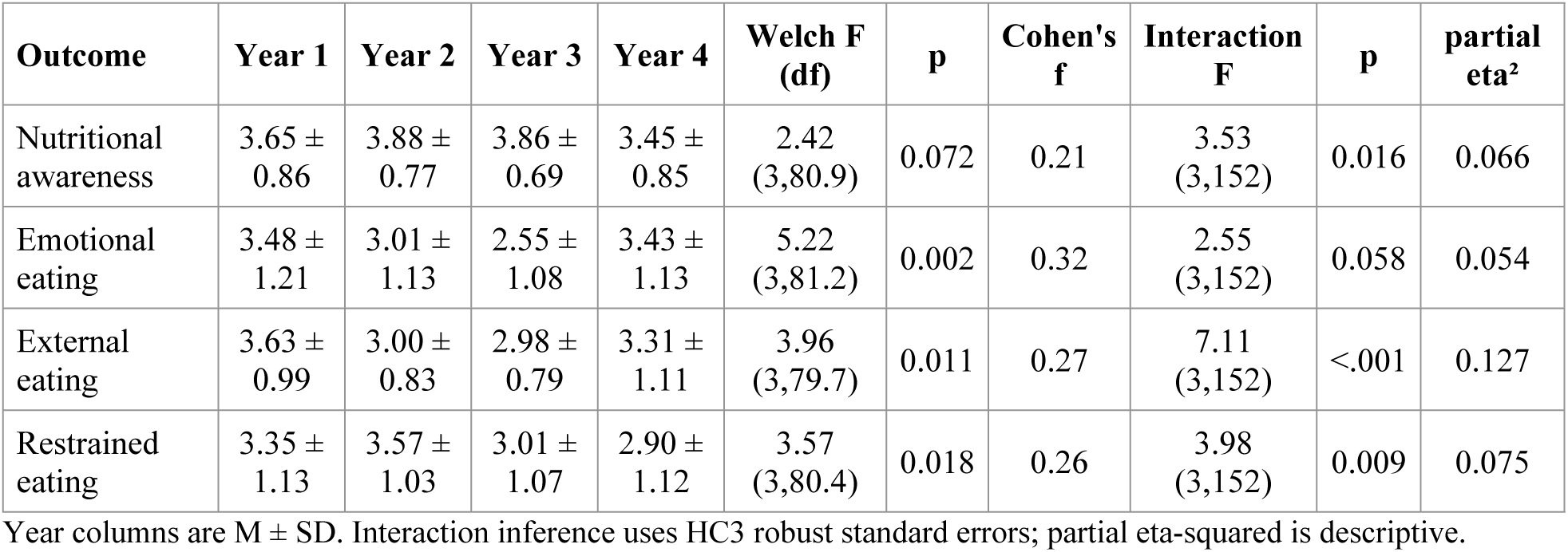
Academic-level effects and sex-by-level interactions.

**Table 10.** Significant Games-Howell comparisons.

| Outcome | Comparison | Mean difference | p |
| --- | --- | --- | --- |
| Emotional eating | Year 1 - Year 3 | 0.935 | 0.0067 |
| Emotional eating | Year 4 - Year 3 | 0.883 | 0.0064 |
| External eating | Year 1 - Year 2 | 0.627 | 0.0159 |
| External eating | Year 1 - Year 3 | 0.649 | 0.0189 |
| Restrained eating | Year 2 - Year 4 | 0.674 | 0.0244 |

*Positive mean differences indicate that the first academic level named had the higher mean.*

The robust factorial models identified sex-by-academic-level interactions for nutritional awareness (HC3 F = 3.53, p = 0.016, partial eta² = 0.066), external eating (F = 7.11, p < 0.001, partial eta² = 0.127), and restrained eating (F = 3.98, p = 0.009, partial eta² = 0.075). The emotional-eating interaction did not reach the robust significance threshold (p = 0.058). The complete HC3-robust interaction tests are summarized in Table 11. Holm-adjusted contrasts showed that fourth-year men reported lower nutritional awareness than fourth-year women (3.09 vs 3.87; adjusted p = 0.017). Men reported more external eating in the first year (4.12 vs 3.19; adjusted p = 0.014) and fourth year (3.94 vs 2.56; adjusted p < 0.001). Men also reported less restrained eating in the first year (2.78 vs 3.87; adjusted p = 0.013) and fourth year (2.33 vs 3.58; adjusted p = 0.002). The significant Holm-adjusted simple sex contrasts are presented in Table 12. The external-eating interaction, the strongest of the three, is shown in Fig 4.

**Fig 4.**
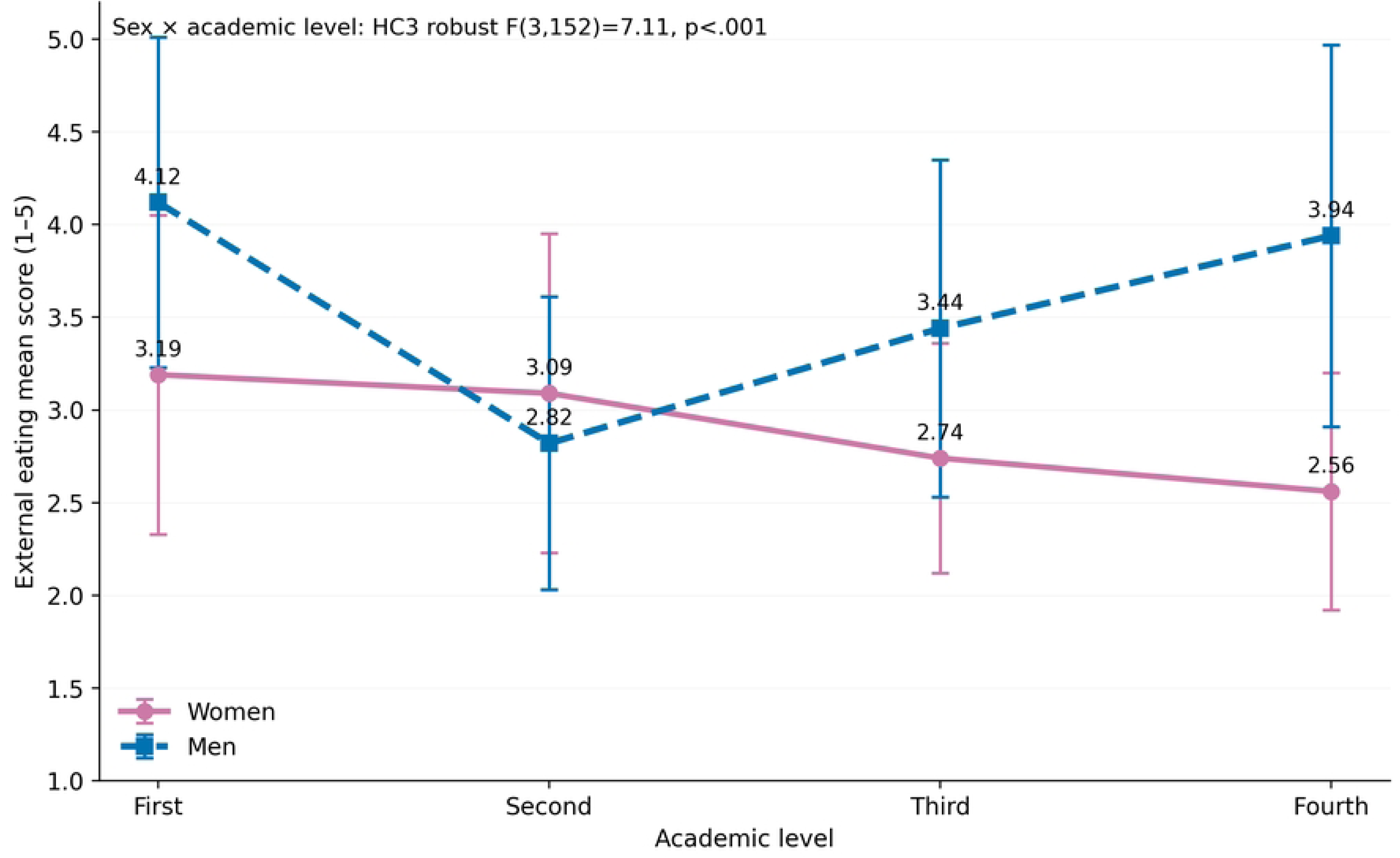
External eating by sex and academic level. Points show means and SD error bars. The crossover pattern corresponds to the strongest sex-by-academic-level interaction.

**Table 11.** HC3-robust sex-by-academic-level interaction tests.

| Outcome | Robust F (df) | p | Partial eta-squared |
| --- | --- | --- | --- |
| Nutritional awareness | 3.53 (3, 152) | 0.016 | 0.066 |
| Emotional eating | 2.55 (3, 152) | 0.058 | 0.054 |
| External eating | 7.11 (3, 152) | <0.001 | 0.127 |
| Restrained eating | 3.98 (3, 152) | 0.009 | 0.075 |

Academic-level cell sizes were: Year 1, 22 women/12 men; Year 2, 26 women/28 men; Year 3, 29 women/6 men; and Year 4, 18 women/19 men. Holm-adjusted simple sex contrasts were used only to interpret statistically significant interactions.

**Table 12.** Significant Holm-adjusted simple sex contrasts.

| Outcome | Academic level | Women M | Men M | Men - women | Holm-adjusted p |
| --- | --- | --- | --- | --- | --- |
| Nutritional awareness | Year 4 | 3.87 | 3.09 | -0.782 | 0.0166 |
| External eating | Year 1 | 3.19 | 4.12 | 0.926 | 0.0140 |
| External eating | Year 4 | 2.56 | 3.94 | 1.381 | 0.00010 |
| Restrained eating | Year 1 | 3.87 | 2.78 | -1.089 | 0.0125 |
| Restrained eating | Year 4 | 3.58 | 2.33 | -1.252 | 0.00217 |

*Inferential conclusions are based on the Holm-adjusted p values. Positive mean differences indicate higher scores among men.*

## Discussion

This study examined nutritional awareness, three eating-behaviour domains, BMI, and equation-estimated RMR in Palestinian physical education students. Several patterns were clear. Nutritional awareness was high, whereas the three eating-behaviour scores were moderate. Awareness was strongly related to all three eating patterns, and these relationships remained after adjustment for basic covariates. BMI and estimated RMR showed much weaker relationships. Sex differences also varied across academic levels, most notably for external eating.

The high nutritional-awareness score does not mean that students necessarily ate well. Food choices at university are shaped by more than knowledge. Time pressure, stress, cost, access to healthy options, social influence, and confidence in cooking can all play a role [1]. Bayomy et al. [3] likewise reported a substantial burden of unhealthy eating habits among Saudi university students, including irregular meals, breakfast skipping, frequent fast-food intake, and inadequate fruit and vegetable intake. Their study assessed eating habits rather than the three Dutch Eating Behavior Questionnaire domains used here, so direct comparison is limited, but it supports examining actual eating behaviour alongside nutrition-related awareness. In another university sample, nutrition literacy was not independently associated with an overall health-promoting lifestyle after adjustment [6]. Nutrition knowledge has also varied by sex, year of study, and faculty [7], while dietary attitudes may be more closely linked with practice than knowledge alone [8]. Palestinian data likewise show associations between nutrition literacy, information seeking, and healthier eating patterns [9]. Regional college research has found poor dietary practices alongside inactivity, sedentary time, and insufficient sleep [17]. Thus, the high awareness score in our sample should be viewed as a favourable starting point rather than proof of an optimal diet. The moderate eating-behaviour scores support this interpretation.

The item-level findings add detail to the high overall awareness score. The most strongly endorsed practices concerned maintaining an appropriate body weight, leaving time between eating and training or practical classes, and consuming a varied diet. The timing result is consistent with evidence linking chrononutrition patterns to diet quality in young adults [35] and with a trial in university rowers in which applied sports-nutrition education improved selected knowledge and dietary intake [10]. By contrast, checking calorie information on food labels was among the least endorsed practices. A 2025 study of 1,153 university students similarly found that knowledge about food labels was high, while routine use of labels and calorie information was less common [36]. This gap suggests that label-reading may be a useful practical target even among students with generally good awareness. The low score for limiting condiments and spices is narrower in meaning and should not be treated as evidence of an otherwise poor diet.

Item responses on the DEBQ also varied within each domain. Boredom was the most prominent emotional-eating cue, whereas fear was the least prominent, indicating that negative emotions were not associated with eating to the same extent. For external eating, visual appeal and taste were the strongest cues. Within restrained eating, guilt after a large intake received the highest mean, while stopping before fullness received the lowest. These patterns reinforce the value of interpreting the three DEBQ domains separately rather than combining them into a single judgement. They also identify practical topics for student support, including boredom-related eating, reactivity to food cues, and concerns about weight control. Population-based data likewise show substantial variation in meal frequency, snacking, timing of hunger, eating speed, eating out, and uncontrolled eating [30]. The guilt item should remain descriptive rather than diagnostic, particularly because Palestinian research has linked body-image dissatisfaction with disordered-eating risk among university women [37].

The clearest behavioural association involved nutritional awareness. Students with higher awareness tended to report less emotional and external eating. One plausible interpretation is that their food choices were less responsive to mood and immediately available food cues. This is compatible with evidence relating stress to emotional eating in college students [25] and with research showing that the university food environment can influence choice [1]. The correlational findings, however, do not determine the direction of these relationships. Stronger self-regulation may contribute to both nutrition awareness and eating control. Sport participation, body image, socioeconomic circumstances, and coaching practices may also influence both variables.241 The adjusted models gave a similar picture to the bivariate correlations. Sex, age, academic level, and BMI together accounted for 12.7%-19.5% of variance in the three eating outcomes. Adding nutritional awareness increased R² by 0.349-0.376, and the adjusted coefficients remained large for emotional eating (β = -0.612), external eating (β = -0.624), and restrained eating (β = 0.636). The observed associations therefore persisted after measured covariates were included. This should not be read as evidence of causation. Adjustment cannot establish temporal order, eliminate unmeasured confounding, or remove the dependence created when predictor and outcome are both self-reported.

Restrained eating should be interpreted differently from emotional and external eating. A higher score denotes a stronger tendency to restrict intake or manage body weight, but it is not a direct measure of diet quality. Among physically active students, some restraint may reflect deliberate meal planning around training or body-mass goals. More pronounced restriction, however, may also occur alongside body-image concerns or poorly regulated eating. Saudi research in female university students has documented marked differences between actual, perceived, and ideal body image [18]. The positive association observed here (r = 0.644) should therefore not be labelled uniformly beneficial. Caution is also supported by Palestinian studies reporting orthorexia-related concerns in physical education students [38] and associations between body-image dissatisfaction and disordered-eating risk among female university students [37]. These findings do not show that restrained eating in our sample was pathological; they indicate the need for future work to distinguish planned dietary control from problematic restriction.

BMI had only small relationships with eating behaviour. After FDR correction, the association with external eating remained significant. The association with emotional eating was borderline (q = 0.051), and restrained eating was unrelated to BMI. University studies have also reported inconsistent relationships between body composition and eating patterns [22]. Regional findings are mixed as well [15,16]. Adult studies provide a broader body-composition context: eating behaviour and sport participation vary across fat-mass and fat-free-mass profiles [27], and dietary or activity patterns relate differently to BMI and body composition across sex and age groups [32]. Among adults with obesity, different eating profiles have also been linked to different body-composition patterns [31]. These populations are not directly comparable with physical education students, but they illustrate a limitation of BMI. It does not describe body composition. The BMI range in our sample was also fairly narrow. Almost two-thirds of students were in the normal category, one quarter were overweight, and none met the obesity threshold. Restricted range may have weakened BMI-behaviour correlations.

RMR must be interpreted carefully because it was estimated rather than measured. The ten Haaf and Weijs equation includes body mass, height, age, and sex [19]. Prediction equations are useful when direct calorimetry is not available, but their error can be important for individual athletes [20]. Resting energy expenditure also depends strongly on body composition and the metabolic activity of different tissues [21]. In a recent study, men had higher basal metabolic rate and lean mass, and eating behaviour also differed by sex and age [29]. BMR and RMR are not identical, but the finding illustrates how metabolic estimates can reflect demographic and body-composition differences. In our sample, the initial correlations of RMR with external and restrained eating were no longer present after adjustment for sex, age, and academic level. This pattern suggests that the unadjusted associations were more likely related to shared demographic characteristics than to resting metabolism itself.

We found sex differences mainly for external and restrained eating. Men scored higher on external eating, whereas women scored higher on restrained eating. The latter pattern agrees with the direction reported in a recent meta-analysis [26]. However, sex differences in eating behaviour vary across populations. Regional studies of adolescents have reported sex-related differences in diet, physical activity, and sedentary behaviour [23]. Large adult studies also describe differences in dietary profiles, body composition, meal habits, and uncontrolled eating [27,28,30,31]. Evidence from university samples is similarly mixed. Associations between stress and maladaptive eating may differ by gender and BMI [39], while DEBQ validation studies have found measurement invariance alongside expected gender differences in scores [14]. Gender has also modified associations involving emotional eating and hedonic hunger [40]. In our study, emotional eating did not differ clearly between men and women. Therefore, patterns reported in other populations should not be assumed to apply unchanged in this setting. A large Italian study published in 2025 also found that weekend eating habits differed by sex and age and were associated with body fat percentage [41].

Academic level contributed additional variation. External eating was highest among first-year students, whereas emotional eating was lowest in the third year. Changes in timetable, independence, and access to food across university life may partly explain these differences [1,2], and nutrition knowledge itself can vary by year of study [7]. The interaction analyses also showed why an overall sex comparison was insufficient. For external eating, men had relatively high means in the first and fourth years, while the means for women were lower at successive academic levels (Fig 4). These between-group patterns should not be interpreted as evidence of within-student change over time. Some sex-by-level groups contained only 12-37 students, so larger samples are needed to test whether the pattern is reproducible.126

The practical implication is not simply to provide more nutrition information. Support is likely to be more useful when it addresses the specific eating pattern reported by the student. For those entering university, priorities may include navigating campus food options, planning meals, managing salient food cues, and using nutrition labels in everyday decisions. Students who eat in response to emotions may also benefit from strategies that address stress and self-regulation. Guidance on restrained eating should clearly separate performance-oriented planning from excessive restriction. Applied sports-nutrition education has improved selected knowledge and dietary intake in university athletes [10], while food-label research shows that positive attitudes do not necessarily translate into regular use [36]. Regional evidence on diet, activity, and overweight also supports multi-behaviour approaches [24]. Reviews of college interventions report improvements in some dietary and physical-activity outcomes, including through digital delivery, although maintenance of these effects remains inconsistent [4,5].

### Study strengths and limitations

Several features strengthen the study. Nutritional awareness, the three DEBQ domains, anthropometric measures, and estimated RMR were assessed in the same group of physical education students. The analysed sample covered 57.8% of the departmental population, and there were no missing values for the main variables. Reliability was high across the study scales. We reported effect sizes and confidence intervals and controlled for multiple testing. Welch procedures and heteroskedasticity-robust methods were used when appropriate. In addition, the key awareness-eating associations were checked in adjusted regression models. We also re-examined the RMR findings after demographic adjustment.

The study has several limitations. The descriptive, analytical, and correlational design identifies relationships and group differences but does not establish temporal order or causation. Both nutritional awareness and eating behaviour were self-reported, leaving the results open to social-desirability and common-method bias. In addition, the awareness measure was drawn from a broader health-awareness scale and focuses on reported practice; its high internal consistency should not be mistaken for evidence that it is an objective test of sports-nutrition knowledge. Future work could pair this measure with more recent instruments for nutrition knowledge or literacy. The Arabic DEBQ also requires further confirmatory testing and formal assessment of measurement invariance in comparable physical education samples. RMR was estimated rather than measured by indirect calorimetry, and BMI cannot distinguish fat mass from fat-free mass, which is particularly relevant in active populations. The sample came from one university and did not include every eligible student. Generalisability beyond this population is therefore limited. Finally, several sex-by-academic-level cells were small, reducing the ability to detect modest interaction effects.

## Conclusions

Among the ANNU physical education students studied, nutritional awareness was high, whereas emotional, external, and restrained eating were moderate. Greater awareness was associated with less emotional and external eating and with more restrained eating, and these relationships remained evident after adjustment for sex, age, academic level, and BMI. BMI and estimated RMR were less consistently related to eating behaviour; the initial RMR associations were no longer evident after demographic adjustment. Men reported more external eating and women more restrained eating overall, but these differences were not uniform across academic levels. The findings favour a targeted approach to nutrition education that addresses stress-related eating, responsiveness to food cues, and weight-control practices. Longitudinal, multi-university studies are needed, ideally using contemporary nutrition measures and direct assessment of RMR, before causal or developmental interpretations are made.

## Data Availability

All relevant data underlying the findings of this study are provided within the manuscript and its Supporting Information files.

## Acknowledgments

The authors thank the students who participated in the study and colleagues in the Department of Sports Sciences at An-Najah National University who supported data collection and instrument review.

